# Increased fibrinaloid microclot counts in platelet-poor plasma are associated with Long COVID

**DOI:** 10.1101/2024.04.04.24305318

**Authors:** Caroline F. Dalton, Madalena IR de Oliveira, Prachi Stafford, Nicholas Peake, Binita Kane, Andrew Higham, Dave Singh, Natalie Jackson, Helen E. Davies, David A. Price, Rae Duncan, Nicola Tattersall, Amanda Barnes, David P. Smith

## Abstract

Outcomes following SARS-CoV-2 infection are variable; whilst the majority of patients recover without serious complications, a subset of patients develop prolonged illness termed Long COVID or post-acute sequelae of SARS-CoV-2 infection (PASC). The pathophysiology underlying Long COVID remains unclear but appears to involve multiple mechanisms including persistent inflammation, coagulopathy, autoimmunity, and organ damage. Studies suggest that microclots, also known as fibrinaloids, play a role in Long COVID. In this context, we developed a method to quantify microclots and investigated the relationship between microclot counts and Long COVID. We show that as a cohort, platelet-poor plasma from Long COVID samples had a higher microclot count compared to control groups but retained a wide distribution of counts. Recent COVID-19 infections were also seen to be associated with microclot counts higher than the control groups and equivalent to the Long COVID cohort, with a subsequent time-dependent reduction of counts. Our findings suggest that microclots could be a potential biomarker of disease and/or a treatment target in some Long COVID patients.

## Introduction

The novel coronavirus SARS-CoV-2 and resulting COVID-19 disease has caused a global pandemic of unprecedented scale and severity. While most patients recover without serious complications, a subset of patients develop prolonged illness, termed Long COVID or post-acute sequelae of SARS-CoV-2 infection (PASC) ^1,2,3,4^. The Office for National Statistics estimates that 2 million people in the UK (3.1% of the population) experience persistent symptoms after acute COVID-19 infection^5^, with over 200 symptoms described, including post-exertional symptom exacerbation (PESE), cognitive dysfunction, dysautonomia and exercise intolerance. Up to 20% of people with a COVID-19 infection suffer for months or years with these symptoms, substantially reducing their ability to carry out day-to-day activities including paid work^6^. There are limited treatments that alleviate the symptoms, with most Long COVID clinics only able to provide support with symptom management. In addition, a small but significant number of people have developed Long COVID-like symptoms after their SARS-CoV-2 vaccination. Castanares-Zapatero *et al* conducted a comprehensive literature review in 2022 of 98 studies^7^ reviewing hypothesis-driven studies and analysing patient data to elucidate Long COVID pathophysiology. They summarised that Long COVID is caused by specific long-term pathologic processes following the initial illness. While acute lung, heart, kidney, liver, and neurological damage can lead to persisting symptoms, additional mechanisms may drive symptoms without clear organ dysfunction. These include autonomic nervous system dysfunction, immune dysregulation causing chronic inflammation and autoimmunity, microvascular dysfunction and hypercoagulation, mitochondrial dysfunction, and occult viral persistence in tissues^8^.

Studies suggest that microclots, also known as fibrinaloids, play a role in Long COVID by blocking capillaries, limiting oxygen exchange, and potentially causing microvascular pulmonary thrombosis and multiple organ failure^9^. Amyloid-containing deposits, resembling microclots, have also been observed in increased numbers in muscle tissue biopsies from people with Long COVID compared to samples from controls^10^, and these numbers increase after exercise, an important observation as many people with Long COVID experience post-exertional symptom exacerbation after activity. However, the deposits in the muscle samples did not appear to be blocking capillaries. Ciceri *et al* coined the term microCLOTS for severe COVID-19 lung injury^11^. They hypothesise initial alveolar damage leads to inflammatory microvascular thrombosis causing ventilation/perfusion mismatch, hypoxemia, and multiple organ failure characteristic of severe COVID-19. Endothelial damage spreading through the microvasculature may drive systemic complications. An alternative hypothesis based on the radiological evidence of lung changes in acute COVID suggests that the oral cavity may provide a direct route for the SARS-COV2 virus to enter the bloodstream and pulmonary vasculature, causing microthombi to form ‘in-situ’ as the primary pathology, with alveolar damage being a secondary phenomenon^12^. This endothelial thrombo-inflammatory syndrome may explain COVID-19’s unique clinical picture.

Kell and Pretorius propose a central role for amyloid fibrinaloid microclots in Long COVID pathophysiology^13^. They previously discovered that fibrinogen can polymerise into protease-resistant "amyloid" fibrin microclots, which are extensively found in Long COVID patient blood samples. Spike protein alone can induce these fibrinaloid microclots, which resist fibrinolysis and are associated with platelet hyperactivation^14^. Supporting this, "triple therapy" anticoagulation treatment with dual antiplatelet therapy (DAPT) plus a direct oral anticoagulant (DOAC) has been reported in a preprint to resolve symptoms in some patients^15,16^.

Fibrinaloid microclots can be induced by adding thrombin *in vitro* to plasma samples from people with various chronic inflammatory and neurodegenerative diseases such as Alzheimer’s, Parkinson’s, type 2 diabetes and rheumatoid arthritis^17,18,19,20^. Following considerable work using electron microscopy (that uncovered ‘dense matted deposits’), it was demonstrated that tiny amounts (1 molecule per 100,000,000 fibrinogen molecules) of bacterial lipopolysaccharide causes blood to clot into an anomalous ‘amyloid’ type form that could be stained with amyloid histological fluorogenic dyes such as thioflavin-T (ThT)^21,22^. These ‘microclots’ are significantly more resistant to breakdown than normal clots, and the phenomena bear similarities to other more classical ‘amyloidosis’^23^. Further work identified extensive microclots in ‘un-induced’ plasma of acute COVID patients^24^ and demonstrated that these could be induced in ‘normal’ blood from uninfected controls simply by the addition of small amounts of purified (virus-free) SARS-CoV-2 (‘alpha’) spike protein^14^.

The initial experiments demonstrating the presence of microclots in the plasma of individuals with Long COVID utilised platelet-poor plasma (PPP) and a blood smear approach^13^. A subsequent method using imaging flow cytometry revealed that samples from people with Long COVID had larger and more frequent microclots than those from controls^25^. The presence of high microclot counts may indicate that a particular treatment, for example, anticoagulant therapy, could succeed in some cases^15,26^. Devloping a robust and objective method to quantify microclot burden in PPP samples will support studies to investigate the efficacy of these therapies in the management of Long COVID and so help to inform treatment.

The primary goal of this study was to investigate the relationship between the microclot count present in PPP, and the occurrence of Long COVID. To determine microclot counts for individuals, we have developed a robust, medium-throughput assay using automated image analysis. In addition, relationships between microclots counts and sex, age, body mass index (BMI), time since COVID-19 infection, and symptoms of Long COVID were investigated. We demonstrate that as a cohort, samples from people with Long COVID have a higher mean microclot count compared to samples from control groups. We also show that samples from people who have recently had a COVID-19 infection have raised microclot counts compared to controls, and that microclot counts decrease over time after a COVID-19 infection.

## Methods

### Participant recruitment and ethical approval

We collected blood samples from four groups all age ≥ 18 years: Long COVID, COVID controls (previously infected), recent COVID (infection within the previous three weeks) and uninfected controls (never knowingly infected and with no active symptoms). The Long COVID group had persistent symptoms lasting at least 12 weeks after a confirmed or probable COVID-19 infection^27^. The COVID control group consisted of individuals who did not report lasting symptoms after a SARS-CoV-2 infection, confirmed by answers to a questionnaire. The recent COVID group consisted of individuals who had experienced a SARS-CoV-2 infection in the past three weeks and had confirmed this via a positive lateral flow or PCR test. We collected data on positive PCR and/or lateral flow tests but did not require a positive test for inclusion as some participants, due to the date of infection, did not have access to these in the early stages of the pandemic but did report symptoms consistent with a COVID-19 infection. Exclusion criteria were current pregnancy, active malignancy or chemotherapy treatment, pre-existing long-term health conditions (assessed by a screening questionnaire). Participants were recruited in Sheffield through word of mouth and social media and in Manchester via the Medicines Evaluation Unit (Manchester University NHS Foundation Trust, Manchester, UK). Ethical approval for the study was obtained from the ethics committees of Sheffield Hallam University (reference number E39973246), and the North West Research Ethics Committee - Preston (reference number 10/H1016/25). All participants provided written informed consent.

### Participant questionnaires

At the time of sample collection, all participants completed online questionnaires hosted and securely stored on Qualtrics. Demographic data collected included sex, age, ethnicity, BMI (Manchester only), information about pre-existing health conditions, dates of COVID-19 infections, dates of positive tests for COVID-19 and dates of SARS-CoV-2 vaccinations. The demographic questionnaire allowed individuals to declare male or female, prefer not to say or would prefer to self-describe, noting that biological sex may differ from the gender the individual identifies with. In this cohort, all participants identified as either male or female. Participants were also provided with a list of common Long COVID symptoms and asked to rate how much they had been affected by that symptom over the previous two weeks.

### Sample collection and processing

Blood was drawn from participants by a qualified phlebotomist into either citrated or EDTA vacutainers. After gentle mixing, whole blood was allowed to stand for 30 minutes and then centrifuged at 3000*xg* for 15 minutes at room temperature. The platelet-poor plasma (PPP) (supernatant) was removed and stored in 150µL aliquots at −80°C until analysis.

### Thioflavin-T staining, image capture and analysis

All sample handling was conducted in a dust-free environment. Aliquots of PPP were defrosted on ice, 40µL of PPP was mixed with 10µL of 100 μM thioflavin-T (ThT) (Merck, Gillingham, U.K.) and incubated for 30 minutes at room temperature protected from light. After incubation 10µL of PPP-ThT was placed in triplicate in µ-Slides 15 Well 3D (Ibidi, Gräfelfing, Germany). Slides were analysed using an Agilent BioTek Cytation 5 cell imaging multimode reader configured with a GFP imaging filter cube and a 4x PL FL objective, controlled by the Agilent BioTek Gen5 microplate reader and imager software. 3×4 images were collected using a 4x objective and the Green Fluorescent Protein (GPF) filter cube with an excitation of 445 to 485nm and emission of 500 to 550nm. Eight z-stacked images were collected with a step size of 53.8μm. Image analysis was performed by combining the z-stack, stitching the images and then background subtraction. Microclots were counted for ThT positive inclusions over a threshold of 2000 arbitrary fluorescence units and between 3 to 100μm in diameter along the longest axis. The analysis resulted in individual counts of microclots for each well; triplicate readings were averaged to give a count for each sample.

### Statistical analysis

Comparative analyses across study groups were conducted employing Kruskal-Wallis tests to ascertain statistical differences, followed by post-hoc evaluations utilising Dunn’s test for pairwise comparisons. Statistical assessments were performed utilising PRISM software.

## Results

The characteristics and demographic information of the participants are reported in Table 1. A higher number of female participants (n=94) were recruited than males (n=35) in all categories. No other differences in demographics were recorded between the control and Long COVID groups, with comparable distributions of age, sex, and BMI. The recent COVID group had a similar age profile to the other groups, but a higher percentage of male participants. BMI data was only available for a subset of participants.

**Table 1.** Participant demographics.

|  | **Long Covid**<br>**(LC)** | **Uninfected**<br>**Controls (UC)** | **Recovered**<br>**Controls (CC)** | **Recent Covid**<br>**(RC)** |
| --- | --- | --- | --- | --- |
| **Number of participants** | 77 | 20 | 32 | 15 |
| **Sex - M/F (%)** | 21/56 (27/73) | 5/15 (20/80) | 9/23 (28/72) | 7/8 (47/53) |
| **Age, years - mean (SD)** | 44.1 (9.9) | 42.2 (14.7) | 41.9 (10.1) | 40.9 (15.1) |
| **\*BMI, kg/m<sup>2</sup> – mean (SD)** | 28.8 (6.9) | 25.2 (3.7) | 25.5 (3.6) | N/A |

**\*BMI data available for 48 Long COVID and 41 control participants**

### Method development

Fibrinaloid microclots are known to consist of amyloid-like material and, hence, can bind to the amyloid-specific dye ThT ^28^. This dye intercalates with the β-sheet structure of amyloid material with a resultant shift in excitation emission and quantum yield. Existing assays have used a blood smear approach whereby PPP is mixed with ThT and incubated before analysing on glass slides^13^. During the development of our method, we identified several processing issues that affected the observed microclot counts with this method. Firstly, we noticed that auto-fluorescent material was visible on untreated glass slides, comparable in size to previously reported microclots. We determined that this material was particulate matter from the laboratory environment or imperfections within the slide’s glass. To counter any false positive results, we established that it was essential to handle all samples in a sterile environment and ensure that all laboratory equipment (tubes, pipette tips, slides) was clean and had not been left outside this environment at any point in the process.

We also identified processing issues relating to the handling and storage of the blood samples. We established that when whole blood tubes were left standing for more than an hour before centrifugation, the resulting PPP showed reduced or no microclot count. After observing this effect, to ensure the accuracy of our analysis, we only used PPP samples that were processed within 30 minutes of collection. Additionally, repeated freeze-thaw cycles of the PPP following processing also resulted in alterations in microclot counts. As such, PPP was stored in aliquots, and analysis was carried out on aliquots that had only been thawed once.

To optimise the efficiency of our process and minimise manual handling, we used 15-well µ-Slides that come pre-sealed and were handled in a dust-free flow cabinet. We also used low protein binding plasticware at all stages. Each PPP sample was imaged directly in the well of the slide, allowing us to obtain 3D images within approximately 10 minutes per well. To minimise variability, we took triplicate technical repeats and analysis parameters were set to automatically identify the boundaries of the well to remove edge effects (Figure 1). Figure 1A shows the captured image, whereas Figure 1B shows the processed image and associated microclot counts, automatically identified by the software. Control experiments and repeat data collection on the same slide gave consistent results up to six hours after sample handling, allowing the use of an autosampler for medium throughput. The researcher who performed the assay was blinded to the analysed PPP category, and each slide contained samples from both the Long COVID and control groups. Our assay provides an unbiased methodology for quantifying microclot counts within PPP samples.

**Figure 1.**
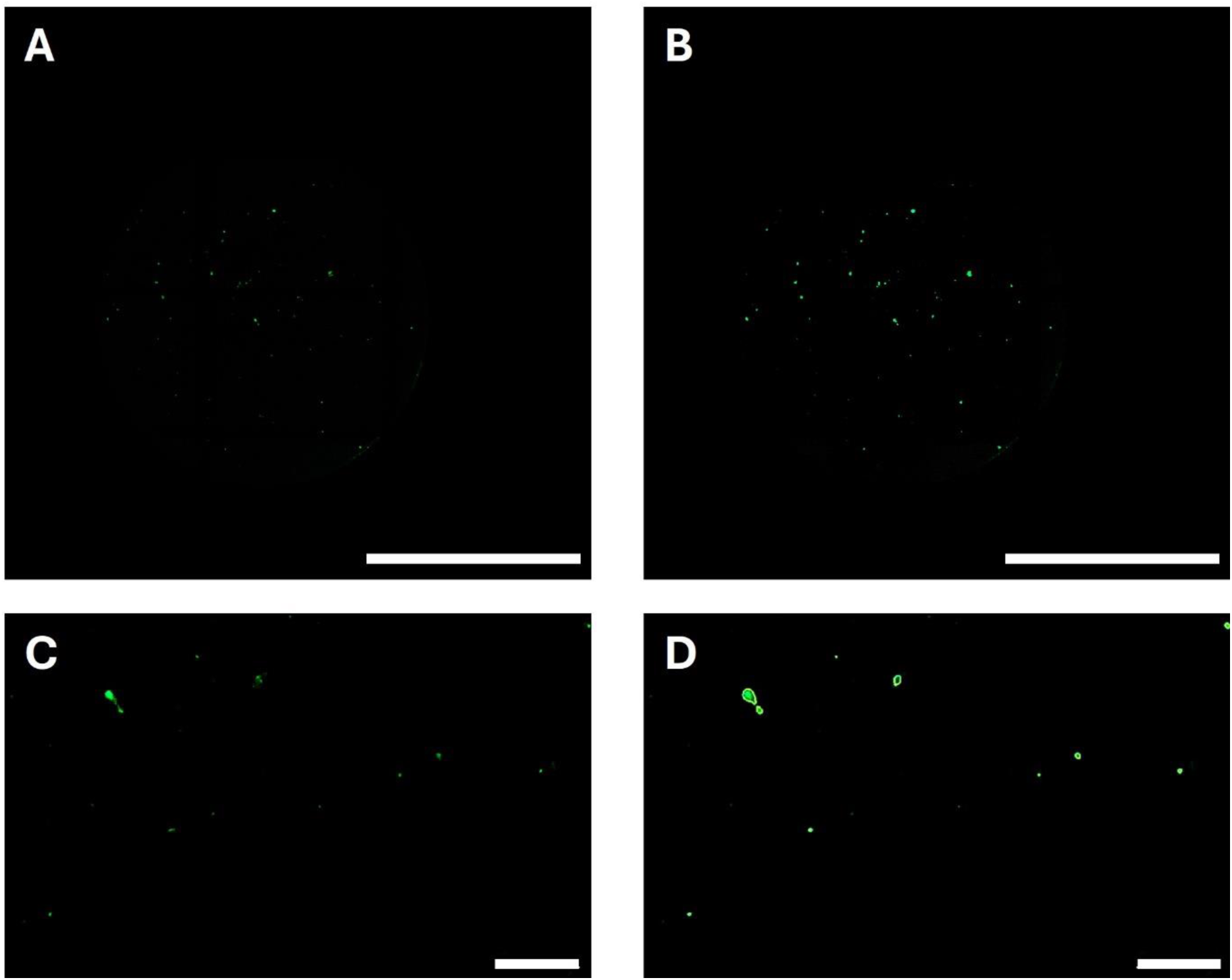
Representative images of microclots in platelet-poor plasma (PPP) stained with Thioflavin-t (ThT). 10µL of PPP-ThT was placed in triplicate in µ-Slides 15 Well 3D slides. Z-stacked images were captured using an Agilent BioTek Cytation 5 cell imaging multimode reader configured with a GFP imaging filter cube and a 4x PL FL objective. Image analysis was performed by combining the z-stacked images, stitching and background subtraction. Microclots were counted for ThT positive inclusions over a threshold of 2000 arbitrary fluorescence units and between 3 to 100µm in diameter along the longest axis. Panels 1A and 1B show z-stacked images before (1A) and after (1B) microclot identification by imaging software, scale bar is 1000µm. Panels 1C and 1D show higher magnification before and after microclot identification, scale bar is 200 µm.

### Microclots in Long COVID samples vs control samples

Having developed a robust assay for the quantification of microclots in PPP samples, we determined the distribution of microclot counts in each of the four groups. Figure 2 shows the average microclot count for each individual. Within the groups, mean microclot counts can be determined for each of the cohorts with the Uninfected Control (mean 13.6 ± S.D. 7.4), COVID Control (20.7 ± 10.1), Long COVID (40.1 ± 28.1) and Recent COVID (50.5 ± 20.4) groups. Significantly higher mean microclot counts were observed in the Long COVID group compared to both uninfected and COVID control groups (*p* < 0.0001 and *p* < 0.01). The control groups also showed a significant difference when compared to the recent COVID group (*p* < 0.0001 and *p* < 0.001). Age did not significantly correlate with microclot counts either as a total grouping or when split by group (Supplementary figure 1). Strikingly, within the data is that only one outlier in the control groups had a microclot count >50 while approximately half the Long COVID group had counts above this level. However, there is overlap between the groups; although microclots are detectable in all samples from people with Long COVID, in about half of the samples the counts are similar to the counts observed in samples from controls.

**Figure 2.**
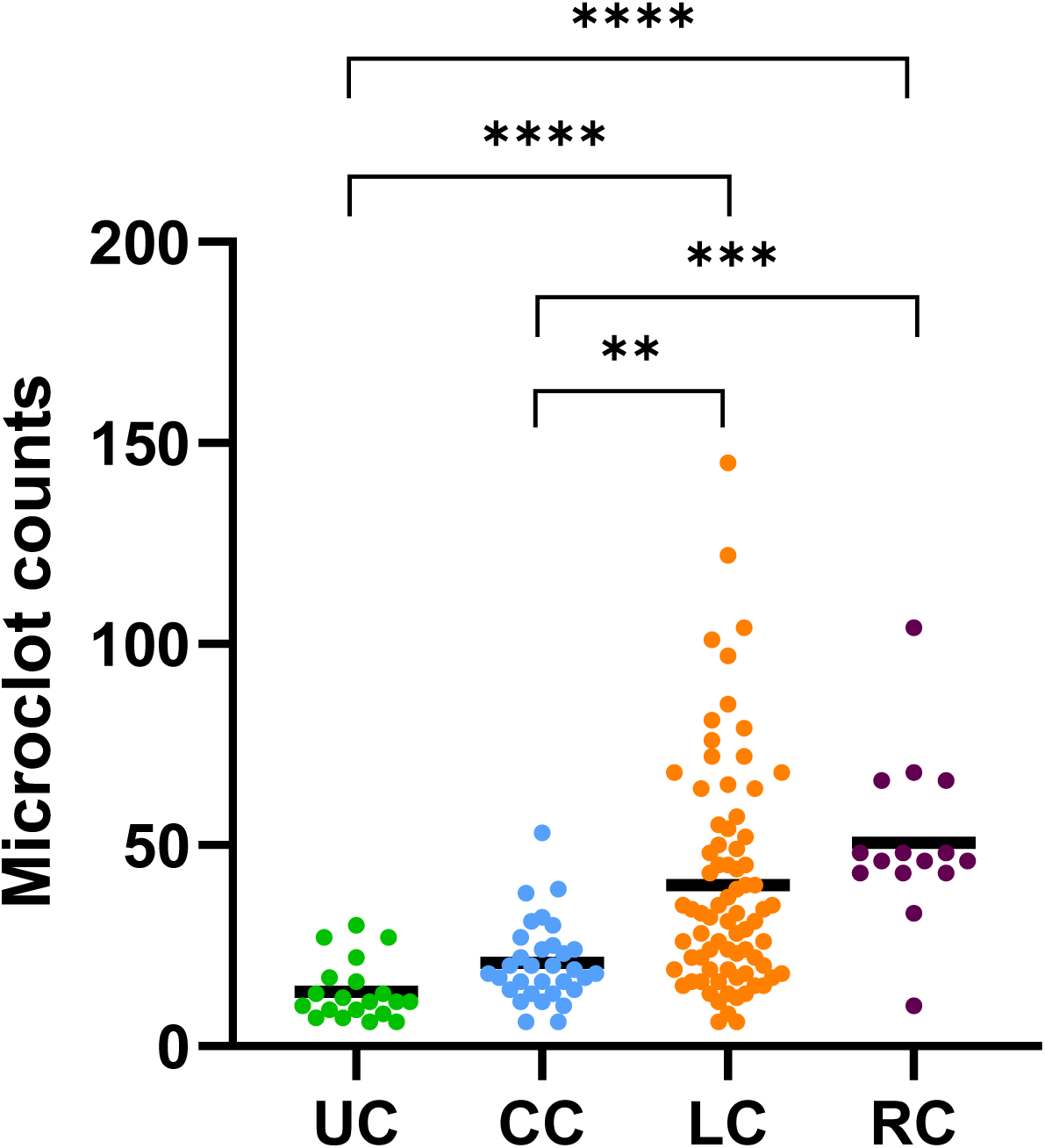
Microclot counts from PPP samples. UC – Uninfected Controls (individuals who have not knowingly been exposed to COVID), CC – COVID Controls (individuals who have had a positive COVID lateral flow or PCR test), LC – Long COVID (individuals who self-identify as suffering from persistent symptoms at least three months after a COVID infection). RC – Recent COVID (individuals who have had a positive COVID lateral flow or PCR test within the previous three weeks). Comparative analyses across study groups were conducted employing the Kruskal-Wallis tests to ascertain statistical differences, followed by post-hoc evaluations utilising Dunn’s test for detailed pairwise comparisons. Statistical assessments was performed utilising PRISM software. Significant differences were seen between the COVID controls and Long COVID individuals (** = *p* < 0.01), between the uninfected controls and Long COVID individuals (**** = *p* < 0.0001) and between both groups of controls and individuals who had tested positive for COVID in the previous three weeks (**** = *p* < 0.0001, *** = p < 0.001).

Pooling the data from the two control groups, Figure 3 shows that female Long COVID participants had a significantly higher mean microclot count than female controls *p* < 0.0001 whereas no significant difference between groups was observed for males.

**Figure 3.**
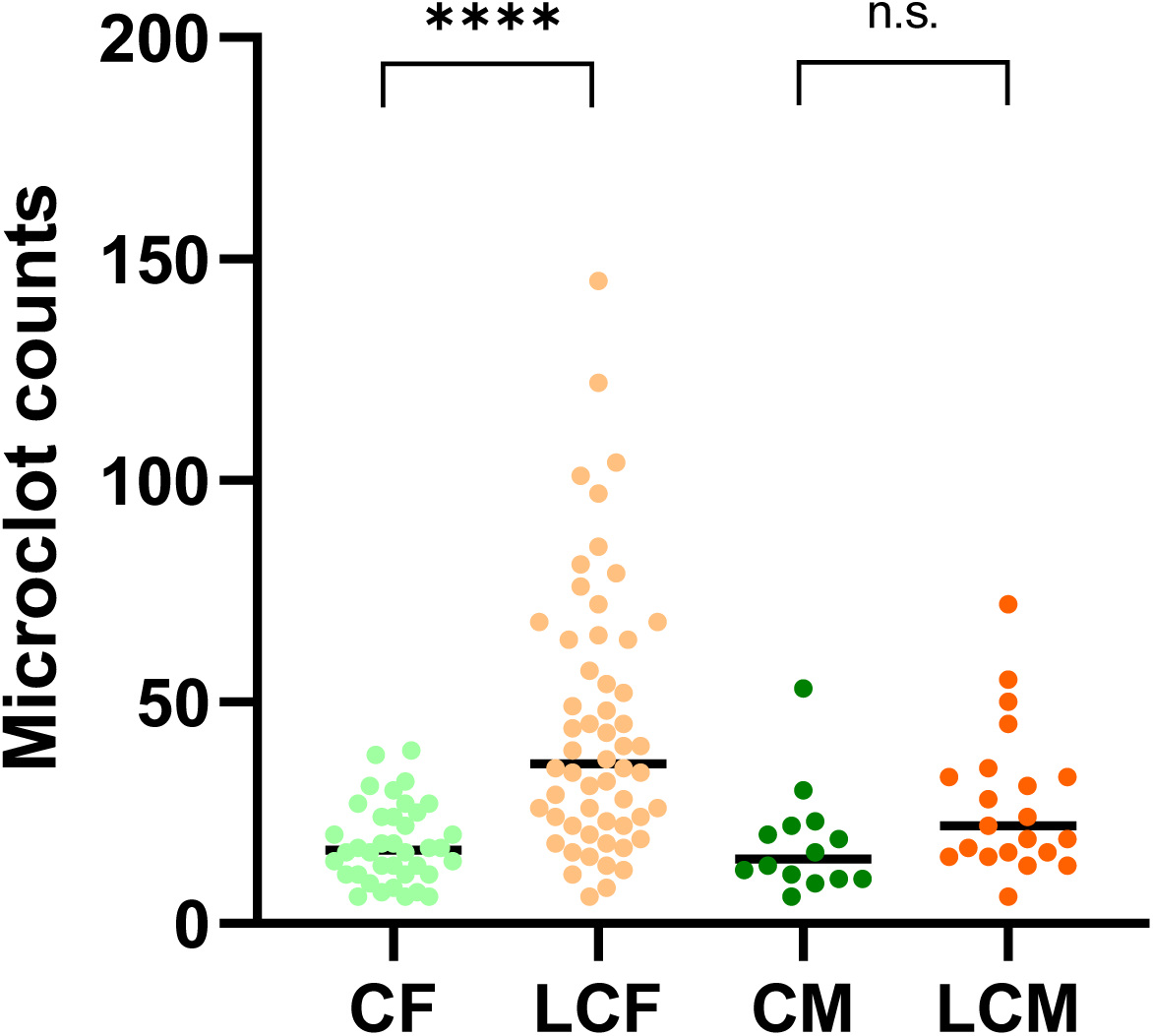
Microclot counts from PPP samples split by sex. CF – Control female, LCF – Long COVID female, CM – Control male, LCM – Long COVID male. Comparative analyses across study groups were conducted employing the Kruskal-Wallis tests to ascertain statistical differences, followed by post-hoc evaluations utilising Dunn’s test for detailed pairwise comparisons. Statistical assessments was performed utilising PRISM software. A significant difference was observed between the female COVID control and the female Long COVID groups (**** = *p* < 0.0001); no significant difference was observed between the male COVID control and the male Long COVID groups.

Recent COVID samples have microclot counts comparable to the upper quartile of the Long COVID group, indicating that recent exposure to the virus leads to higher microclot counts (Figure 2). Within the control group where we have the dates of a confirmed infection from either lateral flow of PCR a significant time dependent decrease in microclot count was observed (Figure 4, r = −0.628, *p* < 0.0001). By ∼450 days post-infection, all samples were within the same range as the COVID control group (20.7 ± 10.1). This indicates that the control group have a decrease in microclot counts to baseline levels within three to four months. Together, these data indicate that exposure to SARS-CoV-2 initially increases the microclot count within PPP, within controls, but these microclots are cleared over time.

**Figure 4.**
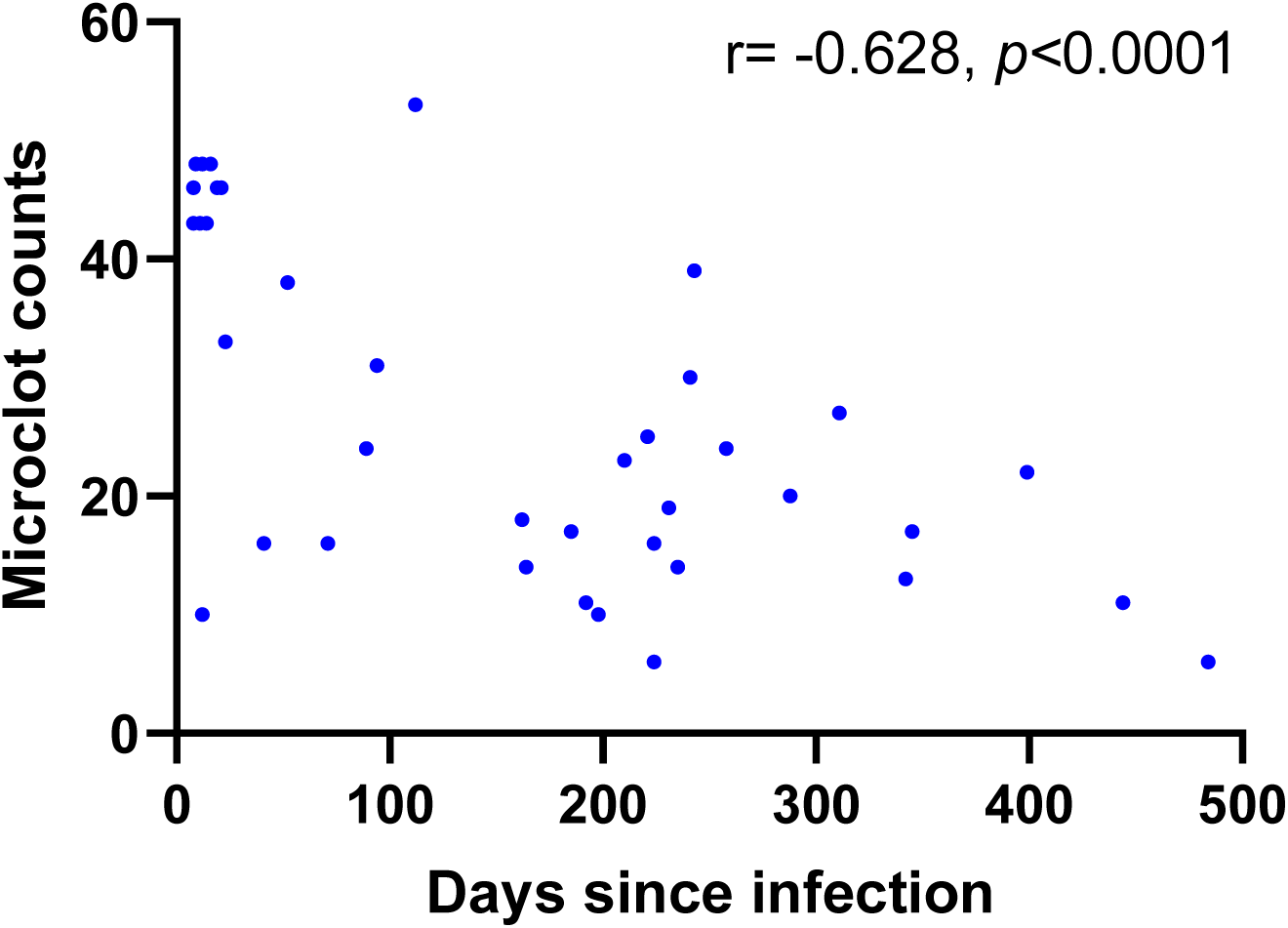
Microclot count as a function of time since infection within the COVID control and recent COVID groups combined. Participants reported the date that they first experienced symptoms during their most recent COVID infection. A Pearson rank correlation showed a significant inverse relationship between microclot count and date of previous COVID infection (r = −0.628 and *p* < 0.0001).

To determine the range of symptoms experienced by the participants with Long COVID each completed an online questionnaire which listed common symptoms associated with Long COVID and other pathologies. All participants were asked to rank if, within the past two weeks, they were “not bothered”, “bothered a little” or “bothered a lot” by symptoms. The list of symptoms was compiled using information from previous studies into Long COVID^6^. This method enabled us to collect information on a wide range of physical symptoms experienced by people with Long COVID using a validated format that has been used in other Long COVID studies^29^, whilst minimising the burden on our participants. The self-reported symptom scores indicate that over 93% of the group experienced "Feeling tired or having low energy", 84% experienced symptoms triggered by physical, mental, or emotional effort, 93% experienced symptoms that occur one or two days after physical, mental, or emotional effort, and 93% experienced difficulty concentrating. These symptoms were experienced at a higher rate than other symptoms (figure 5). These symptoms are typical of post-exertional symptom exacerbation (PESE) and persistent fatigue, which are strongly associated with Long COVID. Using this symptom scoring system we did not see clear relationships between microclot counts and the presence of symptoms. Around half of the participants had microclot counts similar to the controls but reported the same symptom patterns as those with raised microclots, indicating that the presence of elevated microclot counts does not always appear to be a pre-requisite of Long COVID.

**Figure 5.**
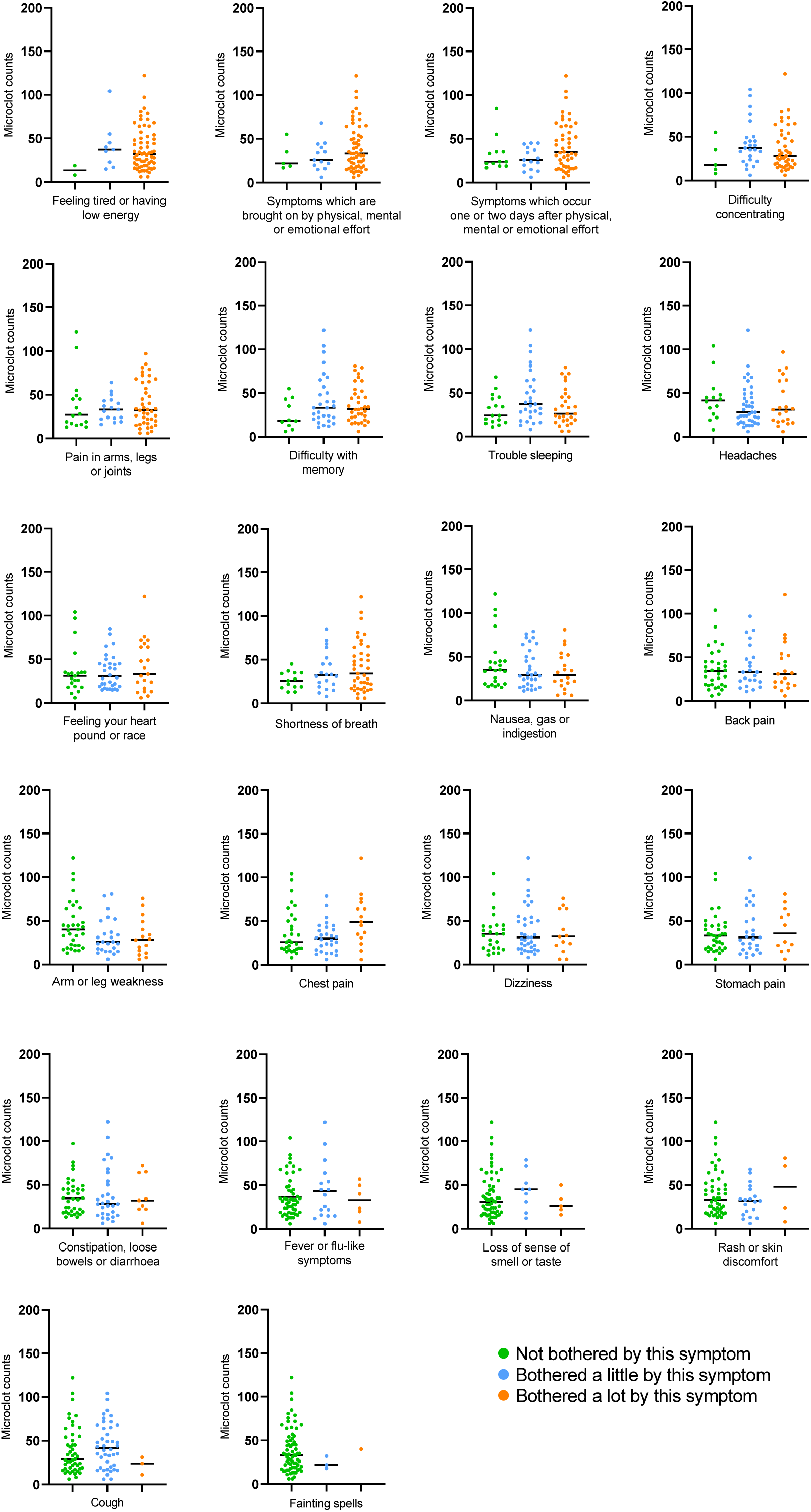
Relationship of microclot counts to symptoms. Long COVID participants rated how much they had been affected by a range of symptoms, and were then split by group based on their response from “not bothered” green, “bothered a little” blue to “bothered a lot” orange. Individuals’ microclot counts were then plotted against these groups. Figures are ordered based on the percentage of participants reporting to be ‘bothered a lot’ by a symptom. Comparative analyses across study groups were conducted employing the Kruskal-Wallis tests to ascertain statistical differences, utilising PRISM software. No clear relationship between microclot counts and degree of symptoms was identified.

## Discussion

Our study aimed to develop a method to investigate the potential role of microclots in Long COVID. We discovered that microclot counts were raised in Long COVID samples compared to controls and that recent COVID-19 cases also had raised counts. However, we also observed that not all people with Long COVID have microclot counts above levels observed in the control samples. The findings suggest that microclots could be a critical factor in Long COVID pathology in a subgroup of people and may also serve as a basis for developing a screening tool to stratify patients for treatment. Our data also show that people with a recent COVID-19 infection have raised microclots, and subsequently take several months to return to control levels. We did not investigate samples from people with other acute viral infections, and it is possible that microclots may be raised in response to other infections. This has implications for other post-viral conditions such as myalgic encephalomyelitis (ME), often associated with infection by Epstein-Barr virus, and we are currently investigating whether people with ME have similar microclot profiles to people with Long COVID.

The potential role of the spike protein in inducing amyloid formation and seeding microclots is intriguing. An *in vitro* study has shown that spike protein spontaneously forms amyloid in the presence of elastase^30^, an enzyme released by activated phagocytes. Cross-seeding between amyloid proteins and peptides has been observed in other situations, and the spike protein amyloid could act to induce micro clot assembly in a similar manner. Proteomics analysis shows complement proteins in microclots^31^, which are known to activate neutrophils and could thereby increase elastase release. Once the initial amyloid seed has formed this may induce aggregation of other molecules, leading to the formation of the microclots. Several studies suggest that SARS-CoV-2 or parts of it, such as the spike protein, potentially remain in body tissues for an extended period., which could trigger persistent microclot formation^10,32^.

Studies indicate an association between Long COVID and both enhanced and diminished immune responses to acute SARS-CoV-2 infection, suggesting a complex relationship between the immune system’s behaviour and Long COVID^33^. Diverse autoantibodies (AAB) specificities have been noted in individuals post-COVID-19, with implications for various bodily systems, including the autonomic nervous system. Our study also found that females had more microclots, which is consistent with the fact that females are more affected by Long COVID^34^. Most Long COVID patients experience severe fatigue, with the female sex group as an independent risk factor in alignment with our symptom data^35^. Female COVID-19 patients exhibit more robust T cell activation, in contrast, male patients have higher levels of innate immune cytokines and a greater presence of non-classical monocytes, potentially accounting for the sex differences in disease outcomes^36^. Males, but not females, who have recovered from COVID-19 show an enhanced response to flu vaccination six months post-recovery, particularly regarding CD8+ T cell-derived IFNγ and a heightened B cell plasmablast and antibody response^37^. This suggests a sex-specific difference in the immune response post-COVID-19, which could have implications for understanding and treating Long COVID. Sex differences in immune response have been attributed to hormones, which could be the primary drivers of this observed difference^38,39^. We also found that microclot counts were not affected by age, but were moderately affected by BMI within the control group which may indicate that raised microclot counts are associated with underlying chronic inflammation.

It is not fully understood how microclotting could relate to the development of Long COVID symptoms. One theory is that the balance of ‘clotting vs anticlotting’ (i.e. fibrin formation vs fibrinolysis) is shifted towards clotting by SARS-COV2 infection. The longitudinal data presented here suggests that this ‘balance’ can be restored within 6 months in those that recover. However, a subgroup of people may have persistent microclots due to ongoing inflammation e.g. due to viral persistence or abnormal immune response, possibly combined with ineffective fibrinolysis, leading to a pro-coagulable state. The aim of treatment with anticoagulant therapy therefore, would be to restore balance to normal coagulation rather than to ‘thin the blood’. Further research into this is required, but measurement of microclots may be a promising biomarker to identify the subgroup of people with Long COVID who may benefit from treatment with anticoagulants.

In addition, previous studies have reported elevated levels of biomarkers such as vascular endothelial growth factor (VEGF), von Willebrand factor (VWF), platelet factor 4 (PF4), serum amyloid A (SAA), α-2 antiplasmin (α-2AP), endothelial-leukocyte adhesion molecule 1 (E-selectin), soluble CD40 ligand (sCD40L) and platelet endothelial cell adhesion molecule (PECAM-1) related to clotting and endothelial dysfunction in Long COVID patients, suggesting thrombotic endothelialitis as a central pathology of the condition^40^. Further studies are necessary to assess levels of these and other markers to determine whether they correlate with microclot counts.

Our study has some limitations that need to be acknowledged. The self-selected group was predominantly white and female, and so is not representative of the full spectrum of people with Long COVID. The differences seen between female and male groups may be partly due to the lower number of samples from male participants. The lack of detailed symptom data limits the analysis of associations between symptoms and microclot counts, we have subsequently incorporated additional symptom questionnaires into our protocols. Since we carried out the sample collection for this study, Long COVID-specific questionnaires have been developed, which facilitate the collection of richer symptom data in people with Long COVID^41^. It is possible some people in the COVID control group may have had some persistent symptoms of SARS-CoV-2 infection that were not captured by our limited symptom questionnaire. These symptoms may have been picked up using the newer Long Covid-specific questionnaires, indicating that these people should be transferred from the COVID control group into the Long COVID group. The uninfected control group was composed of people who reported they had not had a COVID infection, however some of these people may have had undiagnosed asymptomatic infections. Not all the Long COVID patients had positive tests for COVID-19, therefore it is possible in a small set of people that there is an alternative explanation for their symptoms. We also did not control for medication/supplements, which could have affected the results; we recorded this information but did not see a clear pattern relating to microclot counts.

In conclusion, we have developed a robust, automated and quantitative method for determining microclot counts in PPP. However, the role of these microclots requires further research to determine their role in Long COVID pathology and their relationship with COVID-19. Our findings suggest that microclots could be a potential target for treatment in the cohort of Long COVID patients with high microclot counts, indicating the importance of robust phenotyping of this heterogeneous condition. However, further research is necessary to determine the specificity of microclots to COVID-19 and to explore other factors that influence microclot counts.

## Data Availability

All data produced in the present study are available upon reasonable request to the authors

## Acknowledgements

The study team thank all the participants for their contributions to this study, including time, samples and their insight into Long COVID. They are also very grateful for grant funding provided by the Patient-Led Research Fund, a project of Balvi Filantropic Fund (“Balvi”) and Patient-Led Research Collaborative (“PLRC”), and the Open Medicine Foundation for facilitating the funding and administration of the grant.

## Author contributions

CFD, DPS, PS, NP, DAP, HED and RD conceived the study, acquired the funding and supervised the work. Participant recruitment, sample collection and processing were conducted by CFD, MIdeO, BK, AH, DS, NJ and NT. Experimental work was performed by MIdeO. CFD, DPS and AB analysed the data. DPS and CFD drafted the paper. DPS, CFD, PS, NP, BK, RD and DS reviewed and edited the paper. All authors approved the final draft of the manuscript and accept responsibility for the work. AB is an employee of Agilent; and supported the design of the Cytation protocols using blinded data. The other authors have no competing interests to declare.

---

**Supplementary Figure 1.**
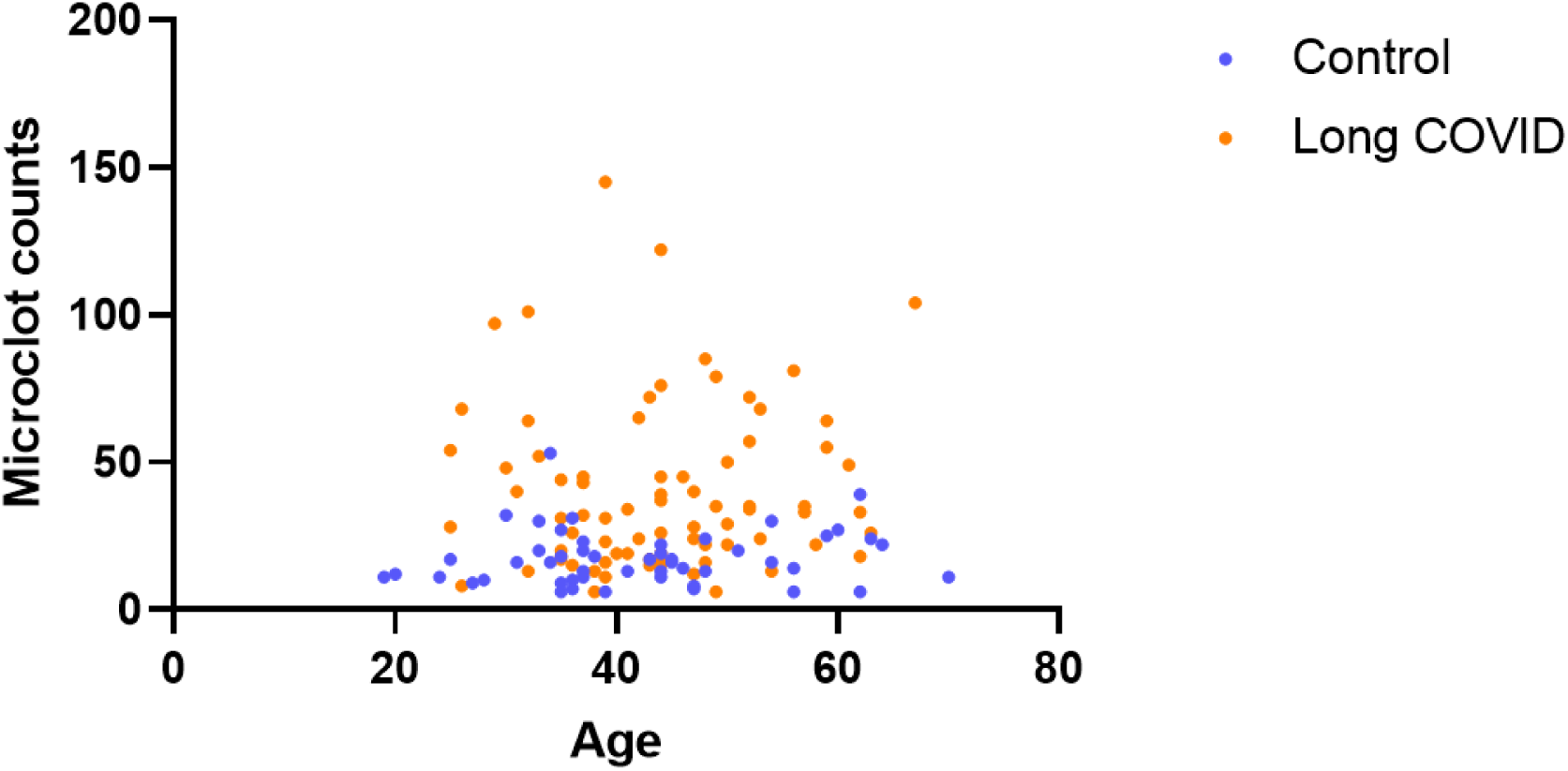
Microclot count was plotted against age in years of the participants for both the COVID control (blue) and Long COVID groups (orange). No significant correlations were observed.

**Supplementary Figure 2.**
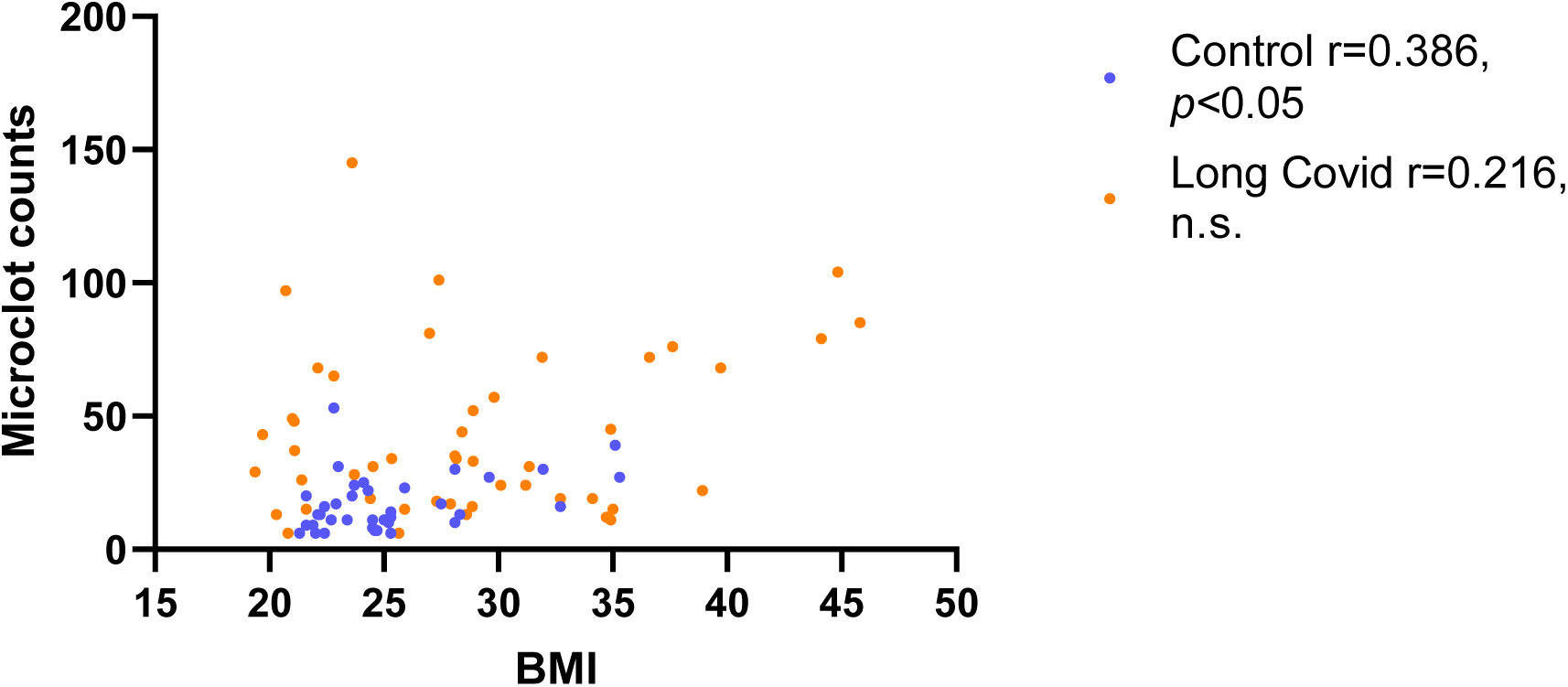
Microclot count was plotted against body mass index (BMI) of the participants for both the COVID control (blue) and Long COVID groups (orange). A Pearson correlation analysis was conducted to examine the relationship between microclot counts and Body Mass Index (BMI) within participants. Specifically, within the COVID control group, the analysis revealed a statistically significant positive correlation between BMI and the number of microclots detected (r = 0.386 and *p* < 0.05).

## References

1 Taquet M, Dercon Q, Luciano S, Geddes JR, Husain M, Harrison PJ. Incidence, co-occurrence, and evolution of long-COVID features: A 6-month retrospective cohort study of 273,618 survivors of COVID-19. PLOS Medicine. 2021;18(9):e1003773. doi: 10.1371/journal.pmed.1003773.

2 Lopez-Leon S, Wegman-Ostrosky T, Perelman C, Sepulveda R, Rebolledo PA, Cuapio A, et al. More than 50 long-term effects of COVID-19: a systematic review and meta-analysis. Sci Rep. 2021;11(1):16144.doi: 10.1038/s41598-021-95565-8.

3 Nalbandian A, Sehgal K, Gupta A, Madhavan MV, McGroder C, Stevens JS, et al. Post-acute COVID-19 syndrome. Nature Medicine. 2021;27(4):601–15. doi: 10.1038/s41591-021-01283-z.

4 Sudre CH, Murray B, Varsavsky T, Graham MS, Penfold RS, Bowyer RC, et al. Attributes and predictors of long COVID. Nat Med. 2021;27(4):626–31. Epub 20210310. doi: 10.1038/s41591-021-01292-y.

5 Prevalence of ongoing symptoms following coronavirus (COVID-19) infection in the UK: 30 March 2023 Office for National Statistics, UK

6 Davis HE, Assaf GS, McCorkell L, Wei H, Low RJ, Re’em Y, Redfield S, Austin JP, Akrami A. Characterizing long COVID in an international cohort: 7 months of symptoms and their impact. EClinicalMedicine. 2021 38:101019. doi: 10.1016/j.eclinm.2021.101019.

7. Castanares-Zapatero, D., Chalon, P., Kohn, L., Dauvrin, M., Detollenaere, J., Maertens de Noordhout, C., Primus-de Jong, C., Cleemput, I., & Van den Heede, K. (2022). Pathophysiology and mechanism of long COVID: a comprehensive review. Annals of medicine, 54(1), 1473–1487. 10.1080/07853890.2022.2076901

8 Davis, H. E., McCorkell, L., Vogel, J. M., & Topol, E. J. (2023). Long COVID: major findings, mechanisms and recommendations. Nature reviews. Microbiology, 21(3), 133–146. 10.1038/s41579-022-00846-2

9 Kell DB, Laubscher GJ, Pretorius E. A central role for amyloid fibrin microclots in long COVID/PASC: origins and therapeutic implications. Biochem J. 2022 Feb 17;479(4):537–559. doi: 10.1042/BCJ20220016.

10 Appelman, B., Charlton, B. T., Goulding, R. P., Kerkhoff, T. J., Breedveld, E. A., Noort, W., Offringa, C., Bloemers, F. W., van Weeghel, M., Schomakers, B. V., Coelho, P., Posthuma, J. J., Aronica, E., Joost Wiersinga, W., van Vugt, M., & Wüst, R. C. I. (2024). Muscle abnormalities worsen after post-exertional malaise in long COVID. Nature communications, 15(1),17.10.1038/s41467-023-44432-3

11. Ciceri, F., Beretta, L., Scandroglio, A. M., Colombo, S., Landoni, G., Ruggeri, A., Peccatori, J., D’Angelo, A., De Cobelli, F., Rovere-Querini, P., Tresoldi, M., Dagna, L., & Zangrillo, A. (2020). Microvascular COVID-19 lung vessels obstructive thromboinflammatory syndrome (MicroCLOTS): an atypical acute respiratory distress syndrome working hypothesis. Critical care and resuscitation : journal of the Australasian Academy of Critical Care Medicine, 22(2), 95–97. 10.51893/2020.2.pov2

12 Lloyd-Jones G, Molayem S, Pontes C C, Chapple I. The COVID-19 Pathway: A proposed oral-vascular-pulmonary route of SARS-CoV-2 infection and the importance of oral healthcare measures. J Oral Med Dent Res 2021; 2: 1–25

13 Pretorius E, Venter C, Laubscher GJ, et al. Prevalence of symptoms, comorbidities, fibrin amyloid microclots and platelet pathology in individuals with Long COVID/Post-Acute Sequelae of COVID-19 (PASC). Cardiovasc Diabetol. 2022;21(1):148. doi:10.1186/s12933-022-01579-5

14 Grobbelaar LM, Venter C, Vlok M, et al. SARS-CoV-2 spike protein S1 induces fibrin(ogen) resistant to fibrinolysis: implications for microclot formation in COVID-19. Biosci Rep. 2021;41(8):BSR20210611. doi:10.1042/BSR20210611

15 Pretorius E, Venter C, Laubscher GJ, et al. Combined triple treatment of fibrin amyloid microclots and platelet pathology in individuals with Long COVID/ Post-Acute Sequelae of COVID-19 (PASC) can resolve their persistent symptoms. Research Square; 2021. DOI: 10.21203/rs.3.rs-1205453/v1.

16 Gert J Laubscher, M Asad Khan, Chantelle Venter et al. Treatment of Long COVID symptoms with triple anticoagulant therapy, 21 March 2023, PREPRINT (Version 1) available at Research Square [10.21203/rs.3.rs-2697680/v1]

17 Pretorius E, Bester J, Page MJ, Kell DB. The Potential of LPS-Binding Protein to Reverse Amyloid Formation in Plasma Fibrin of Individuals With Alzheimer-Type Dementia. Front Aging Neurosci. 2018;10:257. doi:10.3389/fnagi.2018.00257

18 Pretorius E, Page MJ, Mbotwe S, Kell DB (2018) Lipopolysaccharide-binding protein (LBP) can reverse the amyloid state of fibrin seen or induced in Parkinson’s disease. PLoS ONE 13(3): e0192121. 10.1371/journal.pone.0192121

19 Pretorius, E., Page, M.J., Engelbrecht, L. et al. Substantial fibrin amyloidogenesis in type 2 diabetes assessed using amyloid-selective fluorescent stains. Cardiovasc Diabetol 16, 141 (2017). 10.1186/s12933-017-0624-5

20 Pretorius E, Akeredolu OO, Soma P, Kell DB. Major involvement of bacterial components in rheumatoid arthritis and its accompanying oxidative stress, systemic inflammation and hypercoagulability. Exp Biol Med (Maywood). 2017;242(4):355–373. doi:10.1177/1535370216681549

21 Pretorius E, Mbotwe S, Bester J, Robinson CJ, Kell DB. Acute induction of anomalous and amyloidogenic blood clotting by molecular amplification of highly substoichiometric levels of bacterial lipopolysaccharide. J R Soc Interface. 2016;13(122):20160539. doi:10.1098/rsif.2016.0539

22 de Waal GM, Engelbrecht L, Davis T, de Villiers WJS, Kell DB, Pretorius E. Correlative Light-Electron Microscopy detects lipopolysaccharide and its association with fibrin fibres in Parkinson’s Disease, Alzheimer’s Disease and Type 2 Diabetes Mellitus. Sci Rep. 2018;8(1):16798. doi:10.1038/s41598-018-35009-y

23 Kell DB, Pretorius E. Proteins behaving badly. Substoichiometric molecular control and amplification of the initiation and nature of amyloid fibril formation: lessons from and for blood clotting. Prog Biophys Mol Biol. 2017;123:16–41. doi:10.1016/j.pbiomolbio.2016.08.006

24 Pretorius E, Venter C, Laubscher GJ, Lourens PJ, Steenkamp J, Kell DB. Prevalence of readily detected amyloid blood clots in ‘unclotted’ Type 2 Diabetes Mellitus and COVID-19 plasma: a preliminary report. Cardiovasc Diabetol. 2020;19(1):193. doi:10.1186/s12933-020-01165-7

25 Turner, S., Laubscher, G. J., Khan, M. A., Kell, D. B., & Pretorius, E. (2023). Accelerating discovery: A novel flow cytometric method for detecting fibrin(ogen) amyloid microclots using long COVID as a model. Heliyon, 9(9), e19605. 10.1016/j.heliyon.2023.e19605

26 Baker, S. R., Halliday, G., Ząbczyk, M., Alkarithi, G., Macrae, F. L., Undas, A., Hunt, B. J., & Ariëns, R. A. S. (2023). Plasma from patients with pulmonary embolism show aggregates that reduce after anticoagulation. Communications medicine, 3(1), 12. 10.1038/s43856-023-00242-8

27. World Health Organization. (2021). A clinical case definition of post COVID-19 condition by a Delphi consensus, 6 October 2021. World Health Organization. https://iris.who.int/handle/10665/345824

28 Xue, C., Lin, T. Y., Chang, D., & Guo, Z. (2017). Thioflavin T as an amyloid dye: fibril quantification, optimal concentration and effect on aggregation. Royal Society open science, 4(1), 160696.

29 Shen, Q., Joyce, E. E., Ebrahimi, O. V., Didriksen, M., Lovik, A., Sævarsdóttir, K. S., Magnúsdóttir, I., Mikkelsen, D. H., Unnarsdóttir, A. B., Hauksdóttir, A., Hoffart, A., Kähler, A. K., Thórdardóttir, E. B., Eythórsson, E., Frans, E. M., Tómasson, G., Ask, H., Hardardóttir, H., Jakobsdóttir, J., Lehto, K., … Valdimarsdóttir, U. A. (2023). COVID-19 illness severity and 2-year prevalence of physical symptoms: an observational study in Iceland, Sweden, Norway and Denmark. The Lancet regional health. Europe, 35, 100756. 10.1016/j.lanepe.2023.100756

30 Nyström, S., & Hammarström, P. (2022). Amyloidogenesis of SARS-CoV-2 Spike Protein. Journal of the American Chemical Society, 144(20), 8945–8950. 10.1021/jacs.2c03925

31 Pretorius E, Vlok M, Venter C, Bezuidenhout JA, Laubscher GJ, Steenkamp J, Kell DB. Persistent clotting protein pathology in Long COVID/Post-Acute Sequelae of COVID-19 (PASC) is accompanied by increased levels of antiplasmin. Cardiovasc Diabetol. 2021 Aug 23;20(1):172. doi: 10.1186/s12933-021-01359-7.

32 Goh, D., Lim, J. C. T., Fernaíndez, S. B., Joseph, C. R., Edwards, S. G., Neo, Z. W., Lee, J. N., Caballero, S. G., Lau, M. C., & Yeong, J. P. S. (2022). Case report: Persistence of residual antigen and RNA of the SARS-CoV-2 virus in tissues of two patients with long COVID. Frontiers in immunology, 13, 939989. 10.3389/fimmu.2022.939989

33 Altmann, D. M., Whettlock, E. M., Liu, S., Arachchillage, D. J., & Boyton, R. J. (2023). The immunology of long COVID. Nature reviews. Immunology, 23(10), 618–634. 10.1038/s41577-023-00904-7

34 Fernández-de-las-Peñas, C., Martín-Guerrero, J., Pellicer-Valero, O., Navarro-Pardo, E., Gómez-Mayordomo, V., Cuadrado, M., Arias-Navalón, J., Cigarán-Méndez, M., Hernández-Barrera, V., & Arendt-Nielsen, L. (2022). Female Sex Is a Risk Factor Associated with Long-Term Post-COVID Related-Symptoms but Not with COVID-19 Symptoms: The LONG-COVID-EXP-CM Multicenter Study. Journal of Clinical Medicine, 11. 10.3390/jcm11020413.

35 Schulze, H., James, J., Trampe, N., Richter, D., Pakeerathan, T., Siems, N., Ayzenberg, I., Gold, R., & Faissner, S. (2022). Cross-sectional analysis of clinical aspects in patients with long-COVID and post-COVID syndrome. Frontiers in Neurology, 13. 10.3389/fneur.2022.979152.

36 Takahashi, T., Ellingson, M., Wong, P., Israelow, B., Lucas, C., Klein, J., Silva, J., Mao, T., Oh, J., Tokuyama, M., Lu, P., Venkataraman, A., Park, A., Liu, F., Meir, A., Sun, J., Wang, E., Casanovas-Massana, A., Wyllie, A., Vogels, C., Earnest, R., Lapidus, S., Ott, I., Moore, A., Shaw, A., Fournier, J., Odio, C., Farhadian, S., Cruz, C., Grubaugh, N., Schulz, W., Ring, A., Ko, A., Omer, S., & Iwasaki, A. (2020). Sex differences in immune responses that underlie COVID-19 disease outcomes. Nature, 588, 315–320. 10.1038/s41586-020-2700-3.

37 Sparks, R., Lau, W. W., Liu, C., Han, K. L., Vrindten, K. L., Sun, G., Cox, M., Andrews, S. F., Bansal, N., Failla, L. E., Manischewitz, J., Grubbs, G., King, L. R., Koroleva, G., Leimenstoll, S., Snow, L., OP11 Clinical Staff, Chen, J., Tang, J., Mukherjee, A., … Tsang, J. S. (2023). Influenza vaccination reveals sex dimorphic imprints of prior mild COVID-19. Nature, 614(7949), 752–761. 10.1038/s41586-022-05670-5

38. Pollack, B., von Saltza, E., McCorkell, L., Santos, L., Hultman, A., Cohen, A. K., & Soares, L. (2023). Female reproductive health impacts of Long COVID and associated illnesses including ME/CFS, POTS, and connective tissue disorders: a literature review. Frontiers in rehabilitation sciences, 4, 1122673. 10.3389/fresc.2023.1122673

39 Vadakedath S, Kandi V, Mohapatra RK, Pinnelli VBK, Yegurla RR, Shahapur PR, Godishala V, Natesan S, Vora KS, Sharun K, Tiwari R, Bilal M, Dhama K. Immunological aspects and gender bias during respiratory viral infections including novel Coronavirus disease-19 (COVID-19): A scoping review. J Med Virol. 2021 Sep;93(9):5295–5309. doi: 10.1002/jmv.27081.

40 Turner, S., Naidoo, C. A., Usher, T. J., Kruger, A., Venter, C., Laubscher, G. J., Khan, M. A., Kell, D. B., & Pretorius, E. (2024). Increased Levels of Inflammatory and Endothelial Biomarkers in Blood of Long COVID Patients Point to Thrombotic Endothelialitis. Seminars in thrombosis and hemostasis, 50(2), 288–294. 10.1055/s-0043-1769014

41. Hughes, S. E., Haroon, S., Subramanian, A., McMullan, C., Aiyegbusi, O. L., Turner, G. M., Jackson, L., Davies, E. H., Frost, C., McNamara, G., Price, G., Matthews, K., Camaradou, J., Ormerod, J., Walker, A., & Calvert, M. J. (2022). Development and validation of the symptom burden questionnaire for long covid (SBQ-LC): Rasch analysis. BMJ (Clinical research ed.), 377, e070230. 10.1136/bmj-2022-070230

